# Anti-tRNA synthetase syndrome interstitial lung disease: A single center experience

**DOI:** 10.1101/2021.03.01.21252636

**Authors:** Erin M. Wilfong, Jennifer J. Young-Glazer, Bret K. Sohn, Gabriel Schroeder, Narender Annapureddy, Erin A. Gillaspie, April Barnado, Leslie J. Crofford, Rosemarie Beckford Dudenhofer

**Author notes:** Corresponding Author: Erin M. Wilfong, M.D., Ph.D., 1161 21^st^ Ave So., MCN T-1218, Nashville, TN 37232.

## Abstract

**Background:** Recognition of Anti tRNA synthetase (ARS) related interstitial lung disease (ILD) is key to ensuring patients have prompt access to immunosuppressive therapies. The purpose of this retrospective cohort study was to identify factors that may delay recognition of ARS-ILD.

**Methods:** Patients seen at Vanderbilt University Medical Center (VUMC) between 9/17/2017-10/31/2018 were included in this observational cohort. Clinical and laboratory features were obtained via chart abstraction. Kruskal-Wallis ANOVA, Mann-Whitney U, and Fisher’s exact t tests were utilized to determine statistical significance.

**Results:** Patients with ARS were found to have ILD in 51.9% of cases, which was comparable to the frequency of ILD in systemic sclerosis (59.5%). The severity of FVC reduction in ARS (53.2%) was comparable to diffuse cutaneous systemic sclerosis (56.8%, p=0.48) and greater than dermatomyositis (66.9%, p=0.005) or limited cutaneous systemic sclerosis (lcSSc, 71.8%, p=0.005). Frank honeycombing was seen with ARS antibodies but not other myositis autoantibodies. ARS patients were more likely to first present to a pulmonary provider in a tertiary care setting (53.6%), likely due to fewer extrapulmonary manifestations. Only 33% of ARS-ILD were anti-nuclear antibody, rheumatoid factor, or anti-cyclic citrullinated peptide positive. Patients with ARS-ILD had a two-fold longer median time to diagnosis compared to other myositis-ILD patients (11.0 months, IQR 8.5 to 43 months vs. 5.0 months, IQR 3.0 to 9.0 months, p=0.003).

**Conclusions:** ARS patients without prominent extra-pulmonary manifestations are at high risk for not being recognized as having a connective tissue disease related ILD and miscategorized as UIP/IPF without comprehensive serologies.

---

Recognition of idiopathic inflammatory myopathy (IIM) spectrum disease is challenging due phenotypic heterogeneity. While the original Bohan and Peter criteria focused on classic skin rashes and muscle weakness [1], there has been increasing recognition that these clinical classifications oversimplify marked heterogeneity and that myositis autoantibodies are associated with a variety of classical clinical presentations (table 1). Interstitial lung disease is particularly prevalent in the anti-tRNA synthetase syndrome (ARS), which classically presents with the triad of arthritis, interstitial lung disease, and mechanic’s hands. Amongst ARS antibodies, however, the frequency of skin, muscle, and lung disease is variable. Patients with anti-Jo-1 antibodies are more likely to have joint and muscle involvement; non-Jo-1 positive patients are more likely to have vascular and cutaneous involvement [2]. Patients with anti-PL7 and anti-PL12 antibodies have more severe pulmonary manifestations [3].

The diagnosis of IIM-ILD remains a challenge. Presently, the American Thoracic Society recommends that CTD-ILD be ruled out clinically and with basic serologic screening (anti-nuclear antibodies (ANA), rheumatoid factor (RF), and anti-cyclic citrullinated antibodies (CCP)), but does not recommend comprehensive myositis serologies [4]. However, not only are basic serologies more apt to be negative in IIM compared other connective tissue diseases, but IIM patients are also less likely to fulfil clinical connective tissue disease (CTD) classification criteria. For example, nearly all patients with SSc-ILD will have a positive ANA, and the clinical classification criteria for SSc have >95% sensitivity in external validation cohorts [5]. In contrast, fewer than 50% of ARS patients have a positive ANA [6], and external validation of the 2017 American College of Rheumatology and European League Against Rheumatism (ACR/EULAR) classification criteria for idiopathic inflammatory myopathies cohorts has yielded sensitivities as low as 71% [7]. For non-Jo1 associated ARS, the sensitivity of the 2017 ACR/EULAR classification criteria is as low as 25% [8]. This low sensitivity has led to the ACR/EULAR funding the CLASS (classification criteria of antisynthetase syndromes) project to develop improved criteria to diagnose of ARS [9].

The goal of this retrospective cohort study was to (1) evaluate the clinical, radiographic, and serologic features of ARS compared to other patients with IIM and SSc, and (2) gain insights into possible etiologies leading to delayed diagnosis and therapy in the ARS subgroup. We hypothesize that ARS patients will have more subtle clinical and laboratory findings compared to other subsets of IIM-ILD and SSc-ILD despite an equivalent physiologic severity of ILD.

## Patients and Methods

### Patient Identification

Institutional Review Board approval was obtained. Patients were identified from either the Myositis and Scleroderma Treatment Initiative Center (MYSTIC) cohort or the ILD in Autoimmune Inflammatory Myopathy Cohort (IAIMC). Patients with suspected systemic sclerosis, idiopathic inflammatory myopathies, mixed connective tissue disease, or interstitial pneumonia with autoimmune features were eligible for referral to the MYSTIC cohort by their treating provider in the outpatient pulmonary, thoracic surgery, or rheumatology clinic, the inpatient rheumatology or pulmonary consulting service, or by their critical care provider in the intensive care unit (VUMC IRB 141415) from September 20, 2017-December 31, 2019. As MYSTIC is a longitudinal convenience cohort, additional patients were identified through the electronic medical record (VUMC IRB 180672) using a keyword search of the following: “dermatomyositis”, “polymyositis”, “anti-synthetase syndrome”, “systemic sclerosis” in any Vanderbilt Rheumatology or Pulmonology affiliated practice visit between July 1, 2018-October 31, 2018. Exclusion criteria included prior inclusion in the MYSTIC cohort and myositis attributed to medications, infection, or another rheumatologic condition (e.g. mixed connective tissue disease. Diagnoses were verified by review of progress notes, laboratory findings, and scanned documents.

**Table 1.** Clinical manifestations of various myositis specific and associated autoantibodies

|  | Autoantibody | Clinical features | Frequency of organ involvement |  |  |
| --- | --- | --- | --- | --- | --- |
|  |  |  | Skin | Muscle | Lung |
| Anti-tRNA synthetase syndrome | Jo-1 [3, 25] | Classic dermatomyositis with typical rash, muscle weakness, ILD, and mechanic's hands | 30-58% | 69.3-86% | 68%-95% |
|  | PL-7 [3, 25] | Hypomyopathic myositis with frequent arthritis, mechanic's hands, and ILD | 29-56% | 40-81% | >90% |
|  | PL-12 [3, 25] |  | 29-70% | 40-69% | >90% |
|  | OJ [26] |  | 0% | 57% | 100% |
|  | EJ [27-29] |  | 0-75% | 0-100% | 50-100% |
| Other myositis specific autoantibodies | Mi-2 [30] | Classical rash with mild to moderate muscle involvement, ILD uncommon | 90% | 100% | 7% |
|  | NXP-2 [31, 32] | Mild to moderate muscle involvement with myalgia, dysphagia, classical skin rashes, increased malignancy risk, can be associated with ILD | 17-32% | >95% | 7-25% |
| | TIF-1γ [33] | Marked skin involvement often with ulceration, muscle weakness, dysphagia, malignancy | >90% | 63% | 9.1% |
|  | SAE [34] | Classic skin rashes associated with mild to moderate weakness, ILD uncommon | >80% | >95% | 18% |
|  | SRP [35] | Necrotizing myopathy with marked weakness, minimal skin involvement, and infrequent ILD | 3% | >90% | 22% |
|  | MDA5 [36] | Frequent and severe ILD with minimal or no muscle involvement and classically ulcerative skin lesions or palmar papules. | 87.5% | 50% | 50% |
| Overlap syndrome autoantibodies | Ku [37] | Overlap syndrome of systemic sclerosis and inflammatory myositis with mild muscle/skin involvement and variable ILD severity | n.r. | 30% | 43% |
|  | U1RNP [38] | Overlap syndrome of systemic lupus, systemic sclerosis, and inflammatory myositis | 60% | 80% | 45% |
|  | U3RNP [39] | Diffuse systemic sclerosis with increased frequency of inflammatory myopathy | >95% | 25% | 61% |
|  | Pm/Scl [40] | Mild myositis and/or systemic sclerosis features frequently with ILD | 85% | 38% | 61% |
| Abbreviations: ILD=interstitial lung disease |  |  |  |  |  |

\*ILD more common in patients with inflammatory myositis compared to other Ku positive patients

### Clinical phenotyping

Clinical phenotyping was performed by chart abstraction to identify date of symptom onset, skin manifestations (digital ulceration/pitting, mechanic’s hands, Gottron’s sign/papules, heliotrope rash, shawl sign, skin ulcerations), Raynaud’s phenomenon, inflammatory arthritis, pulmonary manifestations (ILD, pulmonary arterial hypertension), myocarditis, muscle weakness, esophageal dysmotility and/or reflux. Serologic data collected included ANA titer and pattern, RF, anti-CCP, and comprehensive myositis serologies (Jo-1, PL-7, PL-12, EJ, OJ, SRP, Mi-2, P155/140, Tif-1γ, SAE1, MDA-5, NXP-2, Ro52, Ro60, Pm/Scl-100, U3RNP, U2RNP through ARUP, Salt Lake, UT). Data on cytoplasmic ANA antibodies was not routinely available. Laboratory values for creatinine kinase (CK) were recorded when available. Patients with dermatomyositis or polymyositis met the 2017 ACR/EULAR classification criteria for probable or definite dermato- or polymyositis [10]. Patient with ARS met the diagnostic criteria proposed by Connors et al. [11] but were not required to meet the 2017 IIM classification criteria. SSc patients met the 2013 classification criteria [12]. When documenting first point of contact at VUMC, patients presenting to thoracic surgery were classified as having presented to a pulmonary provider. Basic demographic data such as age, self-identified gender, and self-identified race/ethnicity were also collected.

### Pulmonary phenotyping

Pulmonary phenotyping was performed using chart abstraction and analysis of computerized tomography (CT) scans, chest x-ray, PFTs, echocardiogram, right heart catheterization (RHC), and pathology reports. Patients were classified as having interstitial lung disease if the radiologist reading the clinical CT scan determined that fibrosis or interstitial lung disease was present on CT scan or chest x-ray if CT was unavailable. The severity of restriction or reduction in DLCO was graded as mild (65-79% predicted), moderate (50-64% predicted) or severe (<50% predicted) using the worst available PFT value. Patients were classified as having pulmonary hypertension if an echocardiogram demonstrated RVSP > 40 mmHg or mean PA pressure on RHC >25 mmHg. RHC was considered gold standard in cases of conflicting data. CT reports were analyzed for mention of honeycombing, bronchiectasis, or ground glass opacities. Radiographic classifications of ILD, e.g. usual interstitial pneumonia (UIP), atypical for UIP, nonspecific interstitial pneumonia (NSIP), and organizing pneumonia (OP) as determined by the reading radiologist was recorded. Patients for whom no CT or PFTs were obtained were assumed to have no or mild ILD [13, 14].

### Statistics

All available cases were analyzed. Bias was minimized through blinding during chart abstractions. Missing data was addressed through pairwise deletion. Categorical variables were analyzed using Fisher’s exact test through the GraphPad Quick Calcs Web site: http://www.graphpad.com/quickcalcs/ (accessed November 2020). Differences in continuous variables were tested using Kruskal-Wallis ANOVA for multiple comparisons followed by Mann-Whitney U tests for comparisons between groups in GraphPad Prism v.7.04. All analyses were two-tailed, and *p values* of less than 0.05 were considered statistically significant. Data is reported as the average ± standard error of the mean unless otherwise noted.

## Results

### Demographics

Demographic characteristics are shown in Table 2. One hundred twenty-five patients were included in the study; one patient was excluded due to conflicting historical reports regarding the presence of ILD and the lack of any PFTs or CT scans. Eighty-four patients (20 DM, 4 PM, 16 ARS, 14 dcSSc, 30 lcSSc) were included from the prospective MYSTIC cohort; 40 unique patients were included from electronic IAIMC cohort (17 DM, 8 PM, 12 ARS, 2 dcSSc, 1 lcSSc). Subjects were predominantly female (77.9% IIM, 78.7% SSc) and Caucasian (81.8% IIM, 78.7% SSc). Median disease duration was estimated from the first report of symptom onset to cohort enrolment and was 5.2 years for IIM versus 9.5 years in SSc (p=0.007).

**Table 2.** Demographics

|  | Idiopathic Inflammatory<br>Myopathies (n=77) | Systemic Sclerosis<br>(n=47) | p value |
| --- | --- | --- | --- |
| **Age (years)<sup>†</sup>** | 57.7(48.4,66.7) | 62.3(52.3,69.0) | .013 |
| **Female Gender** | 60 (77.9%) | 37 (78.7%) | 1.00 |
| **Race** |  |  |  |
| Caucasian | 63 (81.8%) | 37 (78.7%) | 0.82 |
| African American | 11 (14.2%) | 7 (14.9%) | 1.00 |
| Other | 3 (3.9%) | 3 (6.4%) | 0.67 |
| **Disease Duration (years)<sup>†</sup>** | 5.2(2.4,10.4) | 9.5(4.8,14.6) | 0.007 |
| **Known Presence of ILD** | 40 (51.9%) | 28 (59.5%) | 0.46 |
| Radiographic and physiologic evidence of ILD | 33/40 (82.5%) | 20/28 (71.4%) | 0.37 |
| Radiographic evidence of ILD with normal<br>PFTs | 2/40 (5.0%) | 1/28 (3.6%) | 1.00 |
| Physiologic evidence of ILD with normal<br>imaging | 5/40 (12.5%) | 6/28 (21.4%) | 0.34 |
| Historic documentation without objective data | 0/40 | 1/28 (3.6%) | 1.00 |
| **Known Presence of Pulmonary Hypertension** | 8 (20%) | 17 (36.1%) | 0.001 |
| **Pulmonology as First Point of Care at VUMC** | 22 (28.5%) | 13 (27.6%) | 1.00 |
| <sup>†</sup>reported as median (interquartile range) |  |  |  |
| Abbreviations: ILD=interstitial lung disease, PFTs=pulmonary function tests, VUMC=Vanderbilt University Medical Center |  |  |  |

### Clinical phenotypes in IIM

Clinical characteristics of IIM patients are shown in Table 3. Thirty-seven patients (48.1%) were classified as DM, 12 (15.5%) were classified as PM, and 28 (36.4%) were diagnosed as ARS. Subjective proximal muscle weakness was less common in ARS (64.3%) than PM (100%, p=0.002) but not DM (73.0%, p=0.09). There was no difference in the frequency of muscle enzyme elevation in ARS (75.0%) compared to DM (78.3%, p=0.77) or PM (91.7%, p=0.40). Dysphagia or GERD was present in 46.4% of ARS patients, which was not statistically different from DM (29.8%, p=0.20) or PM (66.7%, p=0.31). Inflammatory arthritis was noted in 60.7% of ARS patients, which is higher than either DM (27.0%, p=0.01) or PM (8.3%, p=0.004). By convention, cutaneous manifestations are absent in polymyositis. There was a lower prevalence of Gottron’s sign/papules or heliotrope rash in ARS compared to DM (21.4% v. 89.1%, p=0.0001). There was no difference in prevalence between ARS and DM of mechanic’s hands (43.8% v. 24.3%, p=0.18) or Raynaud’s phenomenon (37.8% v. 35.7%, p=1.0).

**Table 3.** Clinical Phenotype of Idiopathic Inflammatory Myopathies Patients

|  | Associated Serologies |  |  | Cutaneous Manifestations |  | Muscle Involvement |  | Arthritis | Dysphagia<br>or GERD | Raynaud's<br>Phenomenon |
| --- | --- | --- | --- | --- | --- | --- | --- | --- | --- | --- |
|  | ANA | RF | Ro52 | Gottron's or<br>Heliotrope<br>Rash | Mechanic's<br>Hands | Proximal<br>Muscle<br>Weakness | Muscle<br>Enzyme<br>Elevation |  |  |  |
| **By Clinical Phenotype\*** |  |  |  |  |  |  |  |  |  |  |
| Dermatomyositis<br>(n=37) | 25/31<br>(80.6%) | 4/17<br>(23.5%) | 8/12<br>(40.0%) | 33/37<br>(89.1%) | 9/37<br>(24.3%) | 27/37<br>(73.0%) | 29/37<br>(78.3%) | 10/37<br>(27.0%) | 11/37<br>(29.8%) | 14/37<br>(37.8%) |
| Polymyositis<br>(n=12) | 6/10<br>(60.0%) | 2/5<br>(40.0%) | 0/6<br>(0%) | 0/12<br>(0%) | 0/12<br>(0%) | 12/12<br>(100%) | 11/12<br>(91.7%) | 1/12<br>(8.3%) | 8/12<br>(66.7%) | 2/12<br>(16.7%) |
| Anti-Synthetase<br>Syndrome<br>(n=28) | 9/25<br>(36.0%) | 4/21<br>(19.0%) | 14/22<br>(63.6%) | 6/28<br>(21.4%) | 12/28<br>(43.8%) | 18/28<br>(64.3%) | 21/28<br>(75.0%) | 17/28<br>(60.7%) | 13/28<br>(46.4%) | 10/28<br>(35.7%) |
| **By Serology** |  |  |  |  |  |  |  |  |  |  |
| Jo-1<br>(n=14) | 3/11<br>(27.2%) | 2/10<br>(20%) | 5/9<br>(55.6%) | 2/14<br>(14.3%) | 8/14<br>(57.1%) | 12/14<br>(85.6%) | 12/14<br>(85.6%) | 13/14<br>(92.9%) | 7/14<br>(50%) | 6/14<br>(42.9%) |
| PL-7<br>(n=5) | 2/5<br>(40.0%) | 1/5<br>(20.0%) | 3/5<br>(60.0%) | 1/5<br>(20.0%) | 2/5<br>(40.0%) | 2/5<br>(60.0%) | 2/5<br>(40.0%) | 1/5<br>(20.0%) | 3/5<br>(60.0%) | 1/5<br>(20.0%) |
| PL-12<br>(n=6) | 1/5<br>(20.0%) | 1/4<br>(14.3%) | 4/6<br>(42.9%) | 3/6<br>(50.0%) | 1/1<br>(16.7%) | 3/6<br>(50.0%) | 5/6<br>(83.3%) | 1/6<br>(16.7%) | 3/6<br>(50.0%) | 2/6<br>(33.3%) |
| EJ<br>(n=1) | 0/1<br>(0%) | 0/0<br>(0%) | 1/1<br>(100%) | 0/1<br>(0%) | 0/1<br>(0%) | 0/1<br>(0%) | 1/1<br>(100%) | 1/1<br>(100%) | 1/1<br>(100%) | 0/1<br>(0%) |
| OJ<br>(n=2) | 1/2<br>(50.0%) | 0/0<br>(0%) | 1/1<br>(100%) | 0/2<br>(0%) | 1/2<br>(50.0%) | 1/2<br>(50.0%) | 1/2<br>(50.0%) | 1/2<br>(50.0%) | 1/2<br>(50.0%) | 1/2<br>(50.0%) |
| Ku | 2/2 | 0/1 | 2/2 | 1/2 | 0/2 | 1/2 | 2/2 | 0/2 | 1/2 | 0/2 |
| (n=2) | (100%) | (0%) | (100%) | (50.0%) | (0%) | (50.0%) | (100%) | (0%) | (50.0%) | (0%) |
| NXP | 3/3 | 0/2 | 1/2 | 2/3 | 0/3 | 2/3 | 3/3 | 0/3 | 1/3 | 0/3 |
| (n=3) | (100%) | (0%) | (50.0%) | (66.7%) | (0%) | (66.7%) | (100.0%) | (0%) | (33.3%) | (0%_ |
| TIF1g | 1/1 | 0/0 | 0/1 | 1/1 | 0/1 | 1/1 | 1/1 | 0/1 | 1/1 | 0/1 |
| (n=1) | (100%) | (0%) | (0%) | (100%) | (0%) | (100%) | (50.0%) | (0%) | (100%) | (0%) |
| Pm/Scl | 4/4 | 2/2 | 2/4 | 3/4 | 3/4 | 4/4 | 4/4 | 2/4 | 3/4 | 0/4 |
| (n=4) | (100%) | (100%) | (50.0%) | (75.0%) | (75.0%) | (100%) | (100%) | (50.0%) | (75.0%) | (0%) |
| SRP | 0/2 | 0/1 | 0/2 | 0/2 | 0/2 | 2/2 | 2/2 | 0/2 | 2/2 | 0/2 |
| (n=2) | (0%) | (0%) | (0%) | (0%) | (0%) | (100%) | (100%) | (0%) | (100%) | (0%) |
| Mi2 | 5/5 | 0/1 | 1/3 | 5/5 | 2/5 | 5/5 | 5/5 | 1/5 | 1/5 | 3/5 |
| (n=5) | (100%) | (0%) | (33.3%) | (100%) | (40.0%) | (100.0%) | (10.0%) | (20%) | (20%) | (40%) |
| *These are non-overlapping phenotypes. All patients with anti-tRNA synthetase antibodies were categorized as anti-synthetase syndrome. |  |  |  |  |  |  |  |  |  |  |
| Abbreviations: ANA=anti-nuclear antibodies, RF=rheumatoid factor, GERD=gastroesophageal reflux disease |  |  |  |  |  |  |  |  |  |  |

Known interstitial lung disease was present in 51.9% of IIM patients. Chest CT scans were available for all but two IIM-ILD patients. ILD was more frequently seen in ARS (78.5%) than in dermatomyositis (40.5%, p=0.00263) or polymyositis (25.0%, p=0.003). In this cohort, all patients with PL-7, PL-12, or EJ antibodies had interstitial lung disease; 80% of Jo-1 positive patients had ILD. Patients with Mi-2 associated dermatomyositis had the lowest frequency of ILD with only 20.0% having known ILD.

Serologically, only 36% of ARS patients were ANA positive compared to 80.6% of DM patients (p=0.004) and 60% of PM patients (p=0.45). Ro52 positivity was observed with similar frequency in ARS and DM (63.6% v.40.0%, p=1.00) but no PM patient was Ro52 positive. There was no statistically significant difference in the frequency of RF positivity.

### Patients with ARS-ILD frequently present to pulmonary providers at a tertiary center

Of ARS patients, 53.6% presented first to pulmonary providers, which is significantly higher than DM (16.2%, p=0.003) or PM (8.3%, p=0.01). There was no difference in the likelihood of Jo-1 or non-Jo1 associated ARS to present first to a pulmonary provider (60.0% v. 75.0%, p=0.65). Of the 15 patients with ARS-ILD who presented initially to pulmonology, 80% of patients had been seen by outside pulmonologists and 40% by outside rheumatologists. Only one had received the correct diagnosis of ARS-ILD prior to tertiary referral. The time from symptom onset to diagnosis could be abstracted for 21/22 ARS-ILD patients, 11/15 DM-ILD patients, and 3/3 PM-ILD patients. Patients with ARS-ILD had a longer median time to diagnosis (11.0 months, IQR 8.5 to 43 months) than other IIM-ILD subsets (5.0 months, IQR 3.0 to 9.0, p=0.003). (Figure 1) There was a trend towards ARS-ILD patients presenting to pulmonary clinic being less likely to meet IIM classification criteria, but this did not reach significance (66.7% v. 85.8%, p=0.6158).

**Figure 1.**
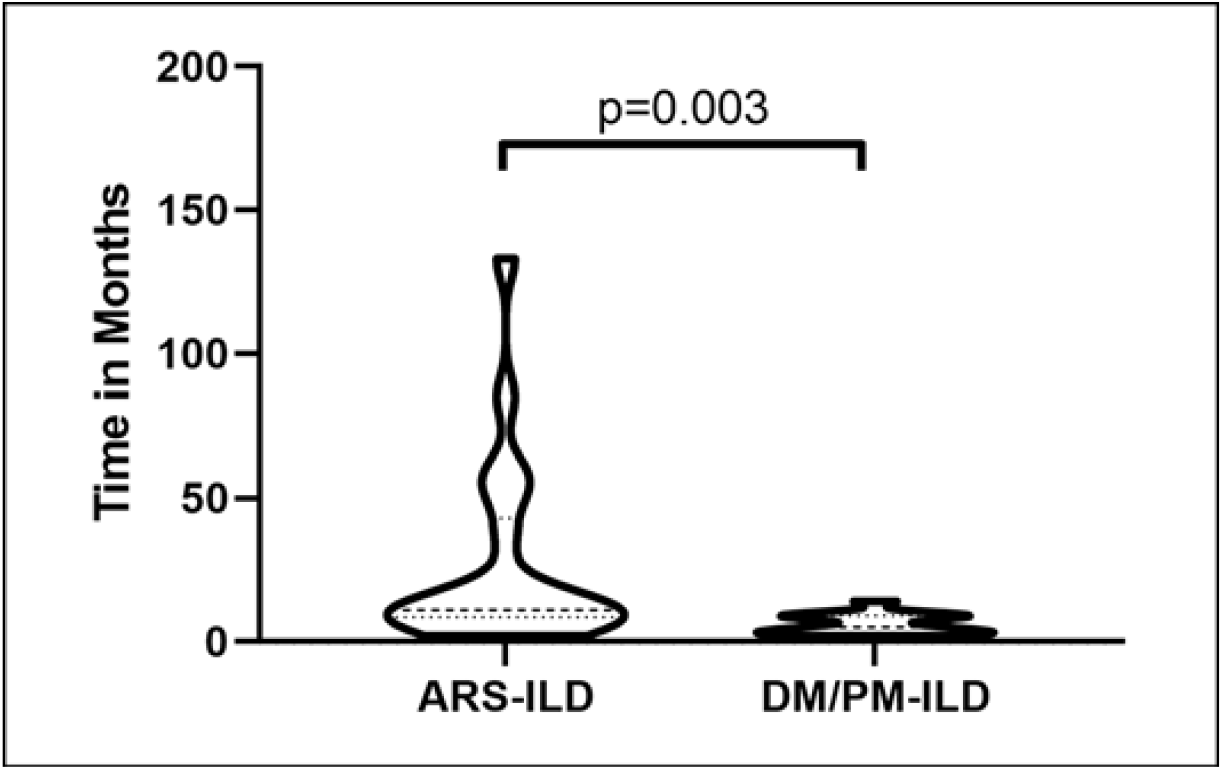
Time to diagnosis for patients with ARS-ILD (n=21) compared to DM/PM-ILD (n=14). Violin plots show the range with interquartile range. Mann-Whitney U-tests shown.

Many pulmonary providers will screen for CTD-ILD using an ANA, RF, and anti-CCP antibodies. The efficacy of detecting a potential CTD-ILD with a basic serologic assessment using ANA, RF, and anti-CCP testing is shown in table 4. Patients with ARS-ILD were much less likely to be identified using this strategy than DM-ILD (33.3% v. 86.7%, p=0.002). If a CK was also obtained, the likelihood of detecting a possible CTD-ILD increased to 72.7% for ARS-ILD compared to 86.7% for DM-ILD (p=0.43). Statistical comparisons to PM-ILD were precluded due to small sample size. Myositis specific serologies were required to detect the final 27.3% of patients with ARS-ILD.

**Table 4.**
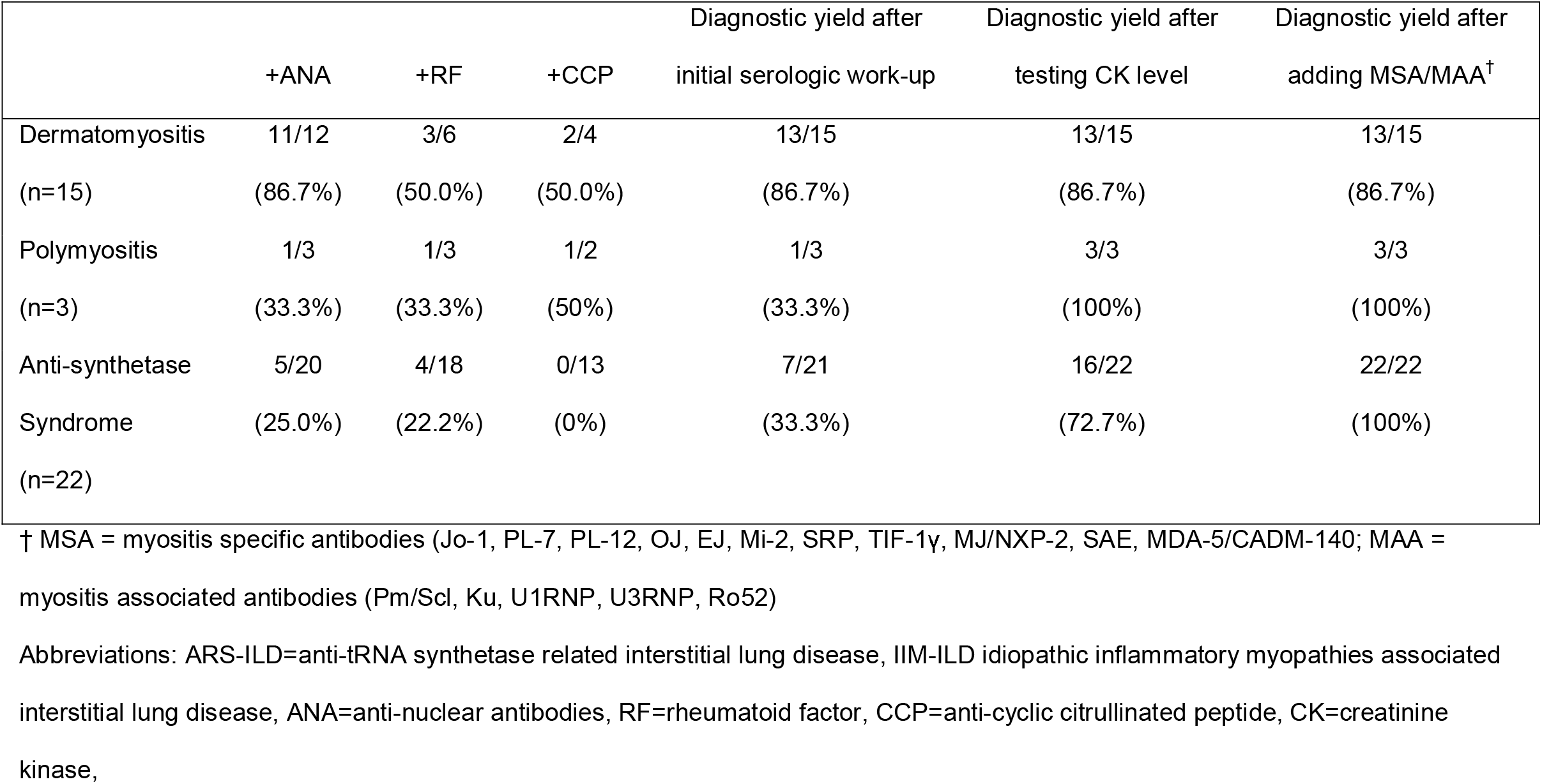
Comprehensive myositis serologies play a critical role in the serologic detection of ARS-ILD compared to other IIM-ILD

|  | +ANA | +RF | +CCP | Diagnostic yield after<br>initial serologic work-up | Diagnostic yield after<br>testing CK level | Diagnostic yield after<br>adding MSA/MAA<sup>†</sup> |
| --- | --- | --- | --- | --- | --- | --- |
| Dermatomyositis<br>(n=15) | 11/12<br>(86.7%) | 3/6<br>(50.0%) | 2/4<br>(50.0%) | 13/15<br>(86.7%) | 13/15<br>(86.7%) | 13/15<br>(86.7%) |
| Polymyositis<br>(n=3) | 1/3<br>(33.3%) | 1/3<br>(33.3%) | 1/2<br>(50%) | 1/3<br>(33.3%) | 3/3<br>(100%) | 3/3<br>(100%) |
| Anti-synthetase<br>Syndrome<br>(n=22) | 5/20<br>(25.0%) | 4/18<br>(22.2%) | 0/13<br>(0%) | 7/21<br>(33.3%) | 16/22<br>(72.7%) | 22/22<br>(100%) |

† MSA = myositis specific antibodies (Jo-1, PL-7, PL-12, OJ, EJ, Mi-2, SRP, TIF-1γ, MJ/NXP-2, SAE, MDA-5/CADM-140; MAA = myositis associated antibodies (Pm/Scl, Ku, U1RNP, U3RNP, Ro52)
Abbreviations: ARS-ILD=anti-tRNA synthetase related interstitial lung disease, IIM-ILD idiopathic inflammatory myopathies associated interstitial lung disease, ANA=anti-nuclear antibodies, RF=rheumatoid factor, CCP=anti-cyclic citrullinated peptide, CK=creatinine kinase,

### Radiographic features of ARS-ILD

Radiographic features of IIM and SSc associated ILD are shown in table 5. Due to the paucity of PM-ILD, these were excluded from statistical comparisons. Notably, only patients with anti-tRNA synthetase antibodies (22.7%) had CT scans that were read by a radiologist as compatible with usual interstitial pneumonia (UIP); no patient a non-anti-tRNA synthetase antibody had a CT scan compatible with UIP. The solitary patient with clinical dermatomyositis and a UIP pattern CT scan did not have comprehensive myositis serologies. Patients with UIP pattern CT scans ranged in age from 49.2 to 70.8 years of age and there was a trend towards the median age of UIP-pattern patients being older than non-UIP pattern patients (63.3, IQR 54.8,70.8 yrs., v. 55.2, IQR 46.8,60.6 yrs., p=0.09). Bronchiectasis trended towards being more prevalent in ARS compared to other IIM subsets (50% v. 22.2%, p=0.10). Of ARS patients, 13.6% had frank honeycombing, which was not seen an any other IIM subset. There was no difference in the frequency of ground glass abnormalities between ARS and other IIM patients.

**Table 5.** Radiographic features of interstitial lung disease in patients with IIM compared to SSc

|  | Radiographic Characteristics |  |  |  |
| --- | --- | --- | --- | --- |
|  | UIP | GGO | BC | HC |
| **Inflammatory Myositis** |  |  |  |  |
| Dermatomyositis | 1/13 | 10/13 | 4/13 | 1/13 |
| (n=15) | (7.7%) | (76.9%) | (30.8%) | (7.7%) |
| Polymyositis | 0/3 | 2/3 | 0/3 | 0/3 |
| (n=3) | (0%) | (66.7%) | (0%) | (0%) |
| Anti-Synthetase | 5/22 | 12/22 | 11/22 | 3/22 |
| Syndrome | (22.7%) | (54.5%) | (50%) | (13.6%) |
| (n=22) |  |  |  |  |
| **By Serology** |  |  |  |  |
| Jo-1 | 2/10 | 9 | 7/10 | 12 |
| (n=10) | (20%) | (90%) | (70.0%) | (20.0%) |
| PL-7 | 1 | 3 | 4 | 3 |
| (n=5) | (20%) | (60%) | (80%) | (60%) |
| PL-12 | 1 | 4 | 4 | 1 |
| (n=5) | (20.0%) | (80%) | (80%) | (20%) |
| EJ | 0 | 0 | 0 | 0 |
| (n=1) | (0%) | (0%) | (0%) | (0%) |
| OJ | 0 | 1 | 0 | 0 |
| (n=1) | (0%) | (100%) | (0%) | (0%) |
| Ku | 0 | 1/2 | 0 | 0 |
| (n=2) | (0%) | (50%) | (0%) | (0%) |
| NXP | 0 | 1 | 1 | 0 |
| (n=1) | (0%) | (100%) | (100%) | (0%) |
| Pm/Scl | 0 | 3/3 | 1/3 | 0 |
| (n=3) | (0%) | (100%) | (33.3%) | (0%) |
| SRP | 0 | 0 | 0 | 0 |
| (n=1) | (0%) | (0%) | (0%) | (0%) |
| Mi2 | 0 | 1 | 1 | 0 |
| (n=1) | (0%) | (100%) | (100%) | (0%) |
| **Systemic Sclerosis** |  |  |  |  |
| lcSSc | 3/15 | 9/15 | 8/15 | 5/15 |
| (n=18) | (20%) | (60%) | (53.3%) | (33.3%) |
| dcSSc | 1/9 | 8/9 | 6/9 | 5/9 |
| (n=10) | (11.1%) | (88.9%) | (66.7%) | (55.6%) |

Abbreviations: IIM=idiopathic inflammatory myopathies,
SSc=systemic sclerosis, UIP = usual interstitial pneumonia (definite or probable), GGO = ground glass opacities, BC = bronchiectasis,
HC = honeycomb changes, lcSSc=limited cutaneous systemic
sclerosis, dcSSc=diffuse cutaneous systemic sclerosis

### Physiologic features of ARS-ILD

Physiologic features of IIM- and SSc-ILD are shown in Figures 2 and 3. Due to the paucity of PM patients with ILD, these were again excluded from subgroup statistical analyses. Overall, patients with IIM and SSc had comparable restriction (60.3% v. 66.6% predicted FVC, p=0.15) and diffusion impairment (48.0% v. 51.1% predicted DLCO, p=0. 63). However, ARS-ILD demonstrated more severe reduction in FVC (53.2% predicted) than patients with DM (66.9% predicted, p=0.005) or lcSSc (71.8% predicted, p=0.005). The severity of ARS and dcSSc-ILD was comparable (53.2% v. 56.8% predicted FVC, p=0.48). Similarly, patients with ARS were more likely to have a severe diffusion impairment (e.g. <50% predicted) than in DM (54.5% v. 6.7%, p=0.004) but not dcSSc (54.5% v. 30.0%, p=0.27). While there was a trend towards worsening impairment of diffusion in ARS relative to DM (39.8% v. 48.0% predicted DLCO, p=0.08) and dcSSc (39.8% v. 51.4% predicted DLCO, p=0.13), this did not reach statistical significance. There was no difference in the severity of restriction or diffusion impairment ARS-ILD patients with positive vs. negative Jo-1 antibodies.

**Figure 2.**
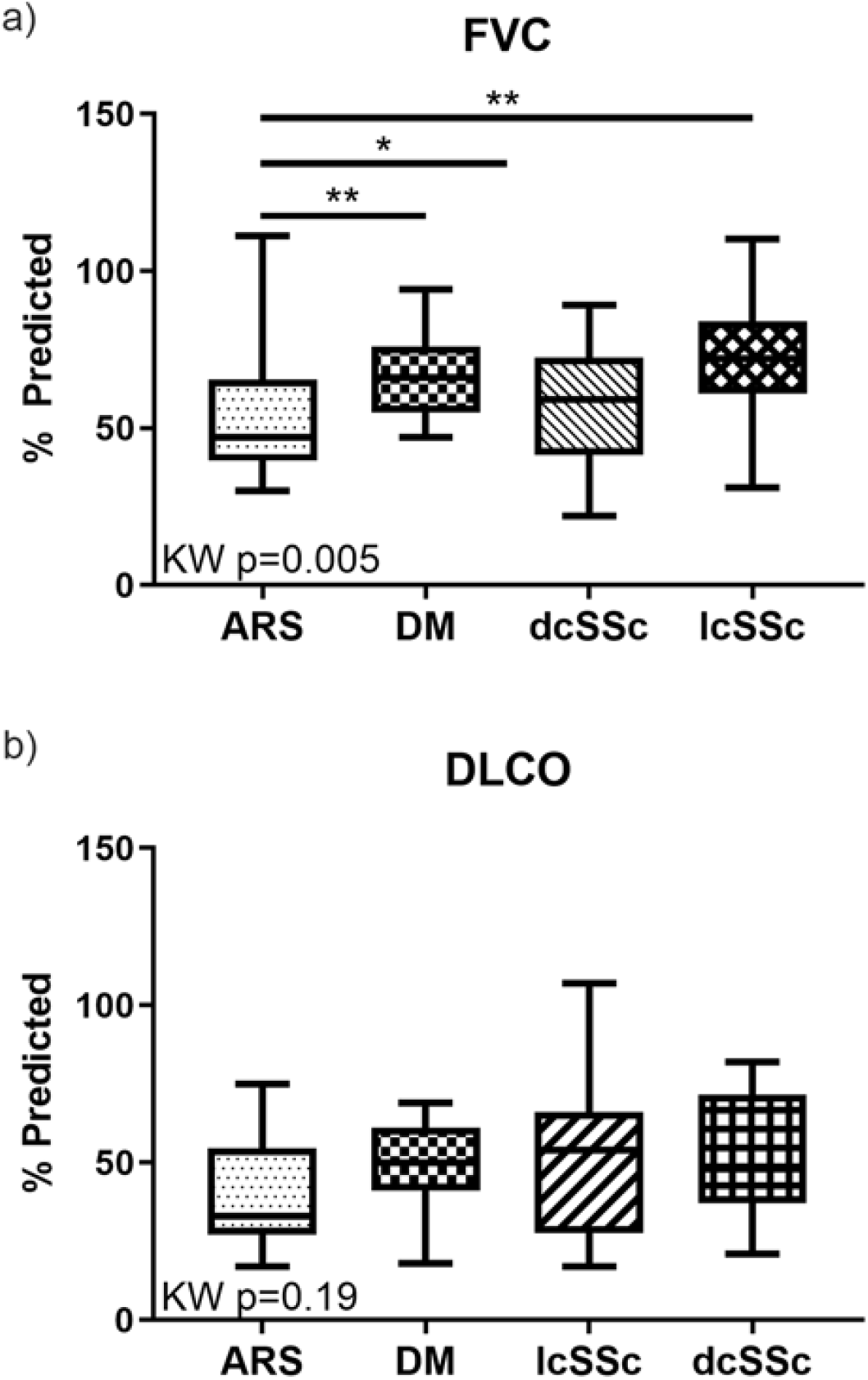
Physiologic features of IIM-ILD comparaed to SSc-ILD. (A) Patients with ARS have more severe restriction than DM or lcSSc and similar restriction compared to dcSSc. (B) There is no significant difference in diffusion impairment amongst IIM or SSc subsets. Box plots depict median ± IQR, KW=Kruskal-Wallis, Mann-Whitney U-test *p<0.05, **p<0.01.

**Figure 3.**
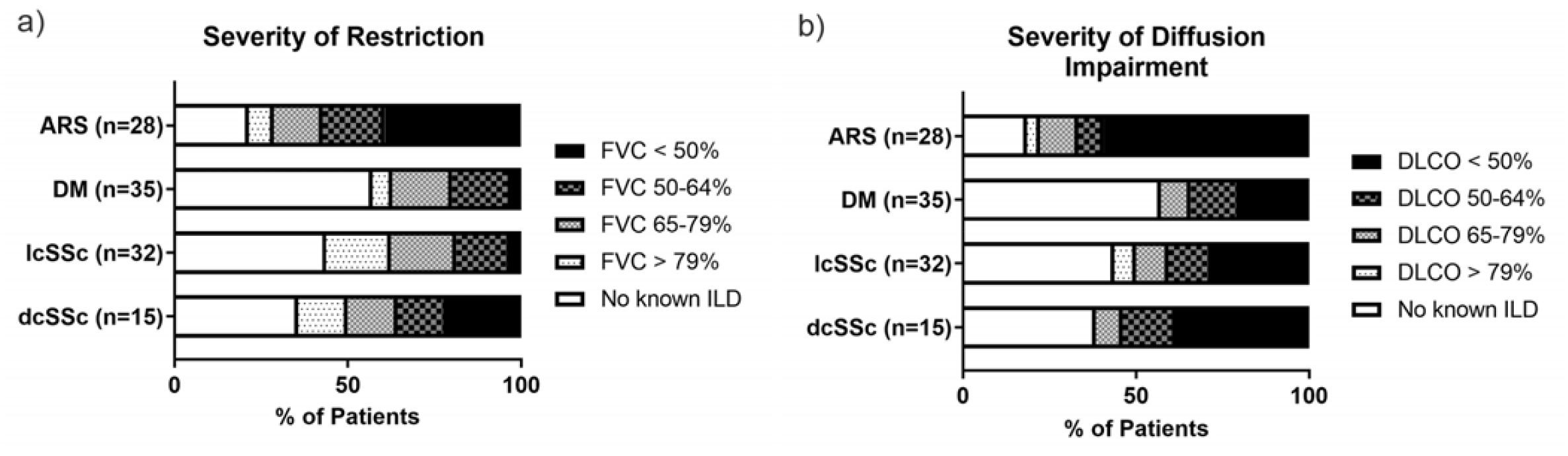
Stratification of physiologic impairment according to American Thoracic Society criteria. The severity of impairment of (A) FVC and (B) DLCO for IIM and SSc subsets are shown.

## Discussion

Our primary findings can be summarized as following, (1) patients with ARS have a high frequency of interstitial lung disease with more severe restriction than other IIM subsets (2) radiographic UIP was observed only in patients with ARS-ILD in this cohort, (3) patients with ARS are frequently referred to a tertiary center after seeing a local pulmonologist and/or rheumatologist without receiving the correct diagnosis, and (4) patients with ARS-ILD have a delayed diagnosis compared to DM-ILD and PM-ILD.

Improving recognition of ARS-ILD is key to ensuring these patients have prompt access to immunosuppressive therapies. Diagnostic delays in ARS are well described and are known to be an independent risk factor for mortality in ARS-ILD [2]. Cavagna et al. described Jo-1 positive patients who presented without the classic triad of arthritis, myositis, and interstitial lung disease had a trend towards increased median time to diagnosis from 2.5 to 10 months [15]. While delays in ARS-ILD diagnosis are frequently discussed in the literature, to our knowledge this is the first report to quantify a delay in diagnosis in ARS-ILD compared to other IIM-ILD. Given that nearly 25% of ARS patients present first with ILD [2], decreasing the diagnostic delay is critical.

The frequency with which ARS-ILD presents with UIP radiographically further complicates recognition of ARS-ILD. In our cohort, 22.7% of patients had CT scans read as compatible with UIP. While ARS-UIP may be enriched at an academic center, it has previously been recognized that most IIM-UIP is associated with ARS. Of the 43 patients in the Pittsburgh cohort with biopsy proven UIP, only one did not have ARS [16]. Using the 2013 Fleischner Society guidelines, a cohort of 69 Chinese ARS-ILD patients identified that 8.7% had a high resolution CT scan classified as UIP [17]. A second longitudinal study of serial CT scans in ARS-ILD found that while 100% of patients had some ground glass at diagnosis, subsequent scans showed the ground glass decreased in 38%. Additionally, 42% of patients had honeycombing and 50% had worsening traction bronchiectasis at follow-up [18]. The prevalence of honeycombing and traction bronchiectasis in ARS may lead to radiographic classification as UIP on CT scans, especially in the community setting. The 2018 American Thoracic Society guidelines for diagnosing idiopathic pulmonary fibrosis allow patients with ground-glass opacities to be classified as UIP in the absence of other clear etiologies and so long as the ground-glass opacities are not the predominant radiographic feature [4]. Thus, ARS patients without prominent extra-pulmonary manifestations are at high risk for not being recognized as having an alternative etiology and miscategorized as UIP/IPF in the absence of comprehensive serologies.

In our cohort, only 33% of patients were identified on basic screening serologies to have signs of a connective tissue disease. While this is slightly lower than some prior studies [6, 17], a substantial number of ARS patients are ANA, RF, and anti-CCP negative in all cohorts. Additionally, there is increasing evidence that obtaining comprehensive serologies and engaging rheumatology improves the care of ILD patients. Nakashima et al. evaluated the ARS serology status of 168 patients with idiopathic interstitial pneumonia and found that 10.7% were positive for an anti-tRNA synthetase antibody, including 5.3% of patients who were diagnosed as IPF [19]. Another study found that even a modest expansion of routine serologies and inclusion of a rheumatologist in the multi-disciplinary ILD team resulted in 21% of IPF patients being re-classified as having an autoimmune process and eight invasive diagnostic procedures were avoided [20]. This study further highlights the importance of using expanded serologies to aid in the identification of IIM-ILD generally and ARS-ILD specifically.

Prompt recognition of ARS-ILD is imperative as immunosuppression is efficacious in ARS-ILD. Previous work by Danoff and co-workers demonstrated that patients with IIM-ILD have an improvement in their FVC following treatment with immunosuppressive therapy [21]. A post-hoc analysis of the rituximab in myositis phase III clinical trial [22] indicated a benefit in ARS-ILD. Efficacy for tacrolimus in both naïve [23] and refractory IIM-ILD [24] has also been reported. Even patients with myositis related usual interstitial pneumonia, which is nearly always ARS-ILD, have a slower rate of FVC decline and improved mortality compared with IPF patients after treatment with immunosuppression [16]. However, if a patient is not recognized to have ARS-ILD, they may not gain access to critically needed immunosuppressive agents. For this reason, we continue to advocate for the use of comprehensive serologies in the evaluation of new patients with interstitial lung disease and the inclusion of rheumatologists as part of the multi-disciplinary team discussion of ILD patients.

This study has several limitations. First, a cross-sectional design at a single center resulted in a limited sample size, which precluded comparisons between some IIM-ILD groups. Additionally, not all CT scans used in this study were high-resolution, which is considered gold standard for the classification of ILD patterns. Further, the use of retrospective chart abstractions meant that not all patients had a complete dataset, and data on objective muscle weakness across the entire clinical course was not available. While comprehensive myositis serologies are clearly useful for identifying ARS-ILD, the yield of comprehensive myositis screening in a tertiary ILD clinic is unclear.

In conclusion, ARS patients without prominent extra-pulmonary manifestations are at high risk for not being recognized as having an alternative etiology and miscategorized as UIP/IPF in the absence of comprehensive serologies. Future work is required to (1) quantify the benefit instituting comprehensive serologies and rheumatologic consultation in the care of ILD patients at a tertiary care center and (2) determine if obtaining comprehensive myositis serologies improves patient outcomes.

## Supporting information

strobe checklist

## Data Availability

All data referred to in this manuscript are included in the main manuscript or supplementary materials.

## Acknowledgements

This work was supported by CTSA award No. UL1TR000445 (EMW) from the National Center for Advancing Translational Sciences, the National Institutes of Health T32HL087738 (EMW), T32AR0590139 (JJY), K08AR072757 (AB), Vanderbilt Faculty Research Scholars Award (EMW), and the Porter Family Fund for Autoimmunity Research. The contents are solely the responsibility of the authors and do not necessarily represent official views of the National Center for Advancing Translational Sciences or the National Institutes of Health.

