## Supplementary material for "Anti-tRNA synthetase syndrome interstitial lung disease: A single center experience": data supplement

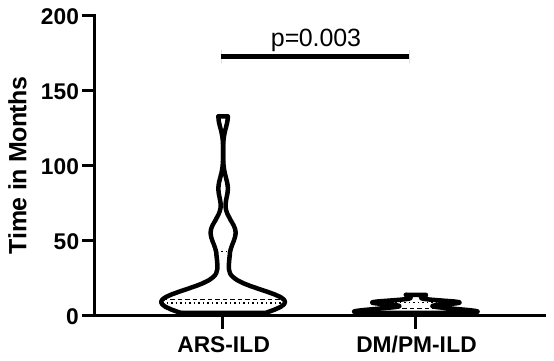

**Figure 1.** Time to diagnosis for patients with ARS-ILD (n=21) compared to DM/PM-ILD (n=14). Violin plots show the range with interquartile range. Mann-Whitney U-tests shown.

| ARS-ILD (time in months to dx) | DM/PM-ILD (time in months to dx) |
| --- | --- |
| 12 | 4 |
| 57 | 14 |
| 7 | 3 |
| 2 | 9 |
| 133 | 10 |
| 13 | 5 |
| 6 | 2 |
| 50 | 3 |
| 9 | 3 |
| 17 | 9 |
| 9 | 9 |
| 36 | 8 |
| 11 | 5 |
| 2 | 2 |
| 9 | 2 |
| 59 |  |
| 12 |  |
| 85 |  |
| 8 |  |
| 11 |  |
| 9 |  |
| Unable to determine |  |

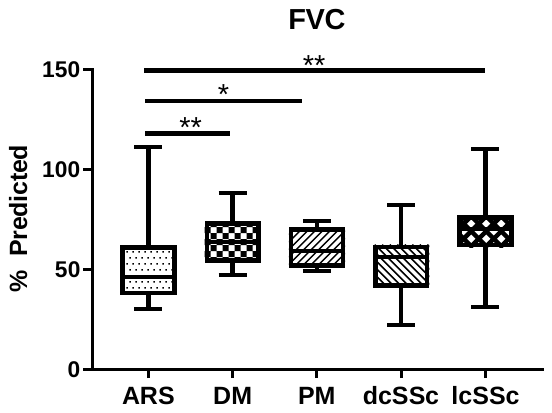

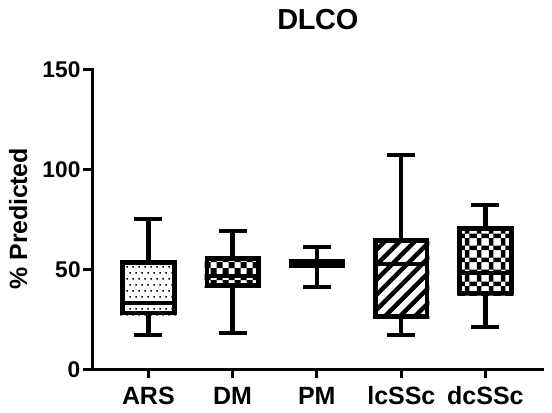

**Figure 2.** Physiologic features of IIM-ILD compared to SSc-ILD. (A) Patients with ARS have more severe restriction than DM or lcSSc and similar restriction compared to dcSSc. (B) There is no significant difference in diffusion impairment amongst IIM or SSc subsets. Box plots depict median ± IQR, KW=Kruskal-Wallis, Mann-Whitney U-test *p<0.05, **p<0.01.

| % Predicted FVC | | | | | % Predicted DLCO | | | | |
| --- | --- | --- | --- | --- | --- | --- | --- | --- | --- |
| ARS | DM | PM | lcSSc | dcSSc | ARS | DM | PM | lcSSc | dcSSc |
| 30 | 54 | 62 | 43 | 76 | 20 | 42 | 41 | 68 | 21 |
| 73 | 74 | 56 | 59 | 84 | 29 | 61 | 61 | 44 | 43 |
| 31 | 76 | 49 | 63 | 61 | 52 | 61 | 53 | 56 | 61 |
| 69 | 51 | 74 | 53 | 70 | 29 | 50 |  | 21 | 53 |
| 62 | 47 |  | 22 | 70 | 33 | 18 |  | 60 | 82 |
| 38 | 66 |  | 60 | 52 | 57 | 69 |  | 23 | 36 |
| 62 | 68 |  | 82 | 31 | 40 | 45 |  | 23 | 40 |
| 54 | 63 |  | 40 | 68 | 17 | 48 |  | 17 | 75 |
| 31 | 76 |  |  | 61 | 75 | 41 |  | 29 |  |
| 62 | 57 |  |  | 56 | 45 | 40 |  | 26 |  |
| 40 | 74 |  |  | 72 | 75 | 32 |  | 54 |  |
| 111 | 88 |  |  | 110 | 23 | 50 |  | 33 |  |
| 43 | 55 |  |  | 77 | 19 | 32 |  | 107 |  |
| 46 | 47 |  |  | 77 | 26 | 43 |  | 65 |  |
| 44 | 53 |  |  | 84 | 30 | 42 |  | 67 |  |
| 53 | 64 |  |  |  | 30 | 63 |  | 51 |  |
| 44 |  |  |  |  | 45 | 55 |  | 88 |  |
| 57 |  |  |  |  | 28 | 51 |  | 57 |  |
| 36 |  |  |  |  | 34 |  |  |  |  |
| 47 |  |  |  |  | 69 |  |  |  |  |
| 34 |  |  |  |  | 60 |  |  |  |  |
